# Understanding Barriers to Diagnosis in a Rare, Genetic Disease: Delays and Errors in Diagnosing Schwannomatosis

**DOI:** 10.1101/2022.01.13.22269170

**Authors:** Vanessa L. Merker, Bronwyn Slobogean, Justin T. Jordan, Shannon Langmead, Mark Meterko, Martin P. Charns, A. Rani Elwy, Jaishri O. Blakeley, Scott R. Plotkin

**Affiliations:** Department of Neurology and Cancer Center, Massachusetts General Hospital, Boston, MA; Department of Neurology and Neurosurgery, Johns Hopkins University, Baltimore, MD; Analytics and Performance Integration, Office of Quality and Patient Safety, Veterans Health Administration, Bedford, MA; Department of Health Law, Policy and Management, Boston University School of Public Health, Boston, MA; Center for Healthcare Organization and Implementation Research (CHOIR), VA Boston Healthcare System, Boston, MA; Center for Healthcare Organization and Implementation Research (CHOIR), VA Bedford Healthcare System, Bedford, MA; Department of Psychiatry and Human Behavior, Brown University Warren Alpert Medical School, Providence, RI

**Author notes:** Corresponding Author: Vanessa L. Merker, PhD, Department of Neurology and Cancer Center, Massachusetts General Hospital, 55 Fruit St. Boston, MA 02114. Grant Numbers: Children’s Tumor Foundation Young Investigator Award 2015-01-005; National Cancer Institute R25CA92203.

**Keywords:** schwannomatosis, rare disease, diagnostic errors, delayed diagnosis, missed diagnosis

## Abstract

Diagnosis of rare, genetic diseases is challenging, but conceptual frameworks of the diagnostic process can be used to guide benchmarking and process improvement initiatives. Using the National Academy of Medicine diagnostic framework, we assessed the extent of, and reasons for diagnostic delays and diagnostic errors in schwannomatosis, a neurogenetic syndrome characterized by nerve sheath tumors and chronic pain. We reviewed the medical records of 97 people with confirmed or probable schwannomatosis seen in two U.S. tertiary care clinics. Time-to-event analysis revealed a median time from first symptom to diagnosis of 16.7 years (95% CI, 7.5-26.0 years) and median time from first medical consultation to diagnosis of 9.8 years (95% CI, 3.5-16.2 years). Factors associated with longer times to diagnosis included initial signs/symptoms that were intermittent, non-specific, or occurred at younger ages (p<0.05). Thirty-six percent of patients experienced a misdiagnosis of their underlying genetic condition (18.6%), pain etiology (16.5%) and/or tumor classification (11.3%). One-fifth (19.6%) of patients had a clear missed opportunity for genetics workup that could have led to an earlier schwannomatosis diagnosis. These results suggest that interventions in clinician education, genetic testing availability, expert review of pathology findings, and automatic triggers for genetics referrals may improve diagnosis of schwannomatosis.

## Introduction

The diagnosis of rare, genetic disorders presents several unique challenges. With more than 9600 recognized rare disorders,^1^ general practice clinicians are unlikely to be familiar with most disorders and often have limited time to access clinical references to aid in diagnosis.

Many genetic conditions are multi-system disorders with clinical manifestations that may be diagnosed and treated by different specialists without consideration of an underlying, unifying genetic diagnosis. Given these diagnostic challenges, it is unsurprising that diagnostic errors and delays are common for people with rare, genetic conditions. In a 2004 survey of approximately 6,000 European patients or caregivers with eight different rare diseases, 41% of respondents reported being previously misdiagnosed and 25% reported a time to diagnosis of ≥5 years.^2^ In a 2013 survey of 887 people with rare diseases and their caregivers in the United States and United Kingdom, respondents experienced an average time to diagnosis of 7.6 years in the U.S. and 5.6 years in the U.K.^3^ These diagnostic errors and delays can have a strong negative impact on patients’ health and quality of life,^4,5^ as well as the health and wellbeing of their family members. Lack of timely genetic counseling can hinder the identification of affected family members and limit patients’ access to family planning resources to reduce the risk of transmitting their disease to offspring.

Comprehensive assessment of the diagnostic process in rare, genetic disorders is thus needed to reduce diagnostic errors, shorten times to diagnosis, and improve health outcomes. In the 2015 landmark report on “Improving Diagnosis in Health Care”, the National Academy of Medicine (NAM) defined diagnostic error as “the failure to a) establish an accurate and timely explanation of the patient’s health problem(s) or b) communicate the explanation to the patient.”^6^ This definition foregrounds patients’ experience of diagnosis – whether and when diagnoses are communicated to the patient, and how delays or misdiagnoses affect patients’ lives. Diagnostic errors can emerge at any point in the cyclical process of gathering, integrating, and interpreting medical information that begins after patients engage in health care. The conceptual model depicted in Figure 1 can be used to highlight the challenges rare disease patients currently face in obtaining timely, accurate diagnoses while also identifying potential areas of clinical practice improvement to help prevent diagnostic errors in the future.

**Figure 1.**
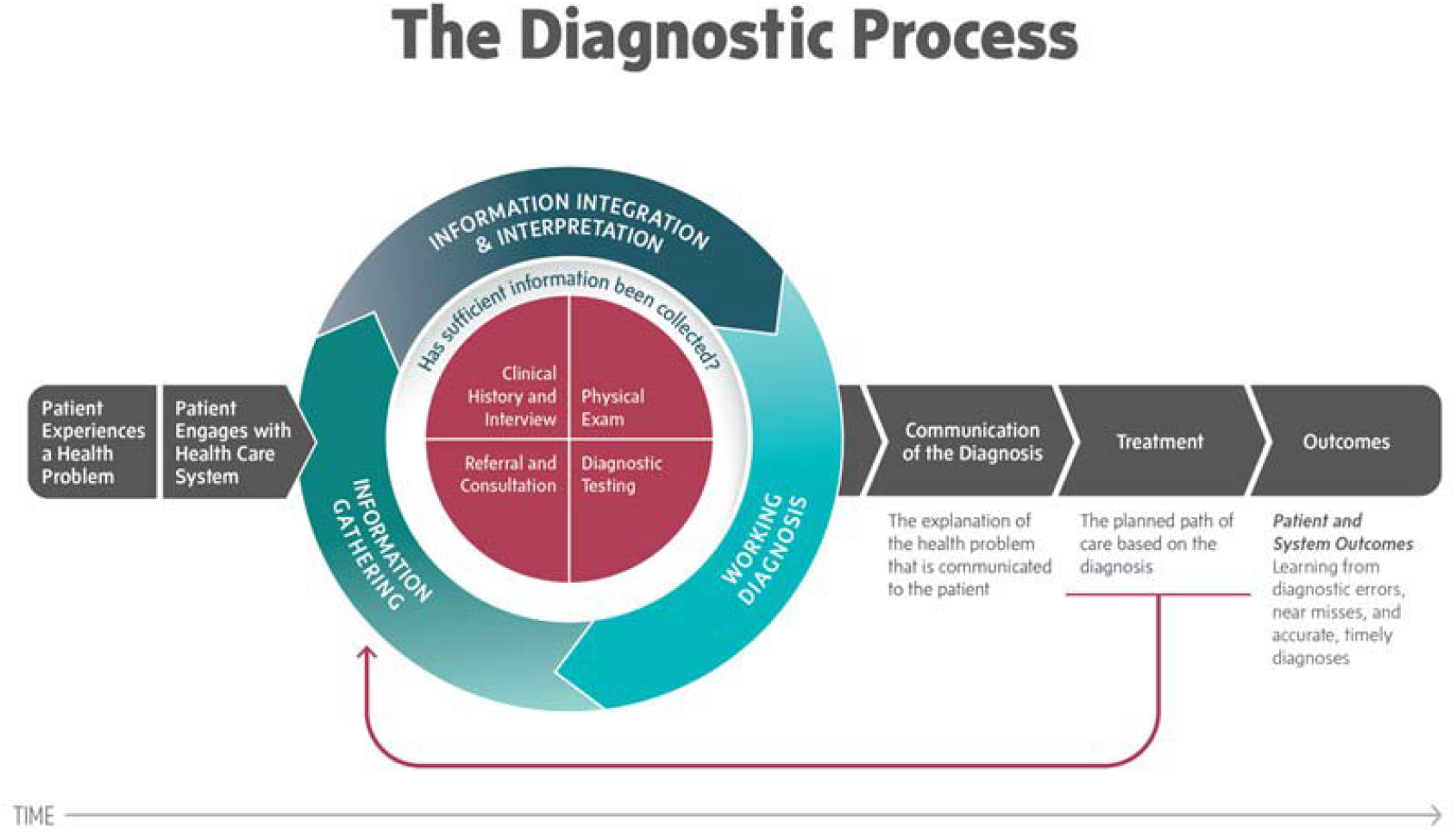
Conceptual Model of the Diagnostic Process (Reprinted with Permission from the National Academy of Medicine)

In this paper, we apply the NAM definition of diagnostic error to study schwannomatosis, a rare, neurogenetic disease characterized by the development of multiple non-malignant nerve sheath tumors and severe, chronic pain.^7^ Multiple features of schwannomatosis make diagnosis challenging. The rarity^8^ and relatively new discovery of schwannomatosis as a distinct entity from other forms of neurofibromatosis (NF)^9,10^ means that many clinicians may be unaware of the condition.^11^ In addition, knowledge of the clinical and genetic features of schwannomatosis is rapidly evolving,^12-15^ and concomitant changes have been made in schwannomatosis diagnostic criteria over the past two decades.^16-18^ Providers may thus be unaware of the current standards for diagnostic ascertainment, newly available genetic testing options, and the full spectrum of symptoms indicative of schwannomatosis. Finally, even when patients are evaluated by expert NF clinicians, phenotypic overlap with other neurogenetic disorders can make diagnoses uncertain.^19^

Previous data on diagnostic delay and misdiagnosis in people with schwannomatosis is limited but suggests errors may occur frequently. In a single-center case series of 87 people with schwannomatosis, the median time from first symptom to diagnosis was 7 years, ranging as high as 39 years.^20^ This case series also reported four patients who had been previously misdiagnosed with malignant tumors. However, this case series did not test for any predictors of diagnostic delay, comprehensively search for prior misdiagnoses, or examine diagnostic communication. To address these limitations, we systematically reviewed the medical records of people with probable or confirmed schwannomatosis eventually seen at two specialized tertiary care clinics. Following the NAM definition of diagnostic error, we assessed the timeliness and accuracy of communication of a diagnosis of schwannomatosis in our cohort by 1) calculating time to diagnosis of schwannomatosis and identifying predictors of diagnostic delay; and 2) determining the rate and types of misdiagnoses of schwannomatosis-related signs and symptoms.

## Methods

### Editorial Policies and Ethical Considerations

All data collection and analysis were conducted in accordance with Institutional Review Board approvals (Partners Human Research Committee Protocol 2014P000633, Johns Hopkins Medicine Protocol 00136913, and Boston University Medical Center Protocol H-32975). A waiver of informed consent was granted for this retrospective medical record review.

### Patient Sample

We performed a retrospective medical record review of people with confirmed or probable schwannomatosis seen at the neurofibromatoses (NF) specialty clinics at Massachusetts General Hospital (MGH) and Johns Hopkins Hospital (JHH). Patients were eligible for inclusion in the study if they 1) had a confirmed diagnosis of schwannomatosis according to the most recent diagnostic criteria^18^ or had a probable diagnosis of schwannomatosis (i.e. did not meet diagnostic criteria but schwannomatosis was considered the leading working diagnosis by their treating physician in the NF clinic), and 2) had at least one in-person visit to the MGH or JHH NF clinics between January 1, 2005 and January 31, 2016. As formal clinical diagnostic criteria for schwannomatosis were not published until 2005, patients diagnosed prior to this year were excluded from the study.^16^

### Data Collection

For eligible patients, we reviewed all inpatient and outpatient records available in the electronic medical record systems of both hospitals through January 31, 2016; for patients seen at MGH, we also reviewed archived paper records when available. The review encompassed provider encounter notes; operative notes; discharge summaries; pathology and imaging reports; and patient communications (such as summaries of phone calls, emails, and patient questionnaires). Records included both internally generated documentation from providers within the MGH and JHH hospital networks as well as externally generated documentation from outside providers that were scanned into electronic patient files or filed into paper charts. Data were abstracted from the medical record by two team members (a researcher and clinician) using a standardized data spreadsheet including the following information:

#### Demographics and Clinical Information

We collected descriptive data on patients’ sex, age, inheritance pattern (sporadic or familial), extent of disease (generalized vs. anatomically limited^16^), type and frequency of initial signs/symptoms of schwannomatosis, initial imaging results (single vs. multiple tumors present), and the specialties of healthcare professionals they consulted.

#### Key Diagnostic Timepoints

We recorded dates for the following key timepoints in the diagnostic process shown in Figure 2: 1) when the patient first experienced a health problem (i.e. a bodily change, sign, or symptom)^21^ related to schwannomatosis, 2) when the patient first had a medical appointment for a health problem related to schwannomatosis; 3) when a diagnosis of schwannomatosis was confirmed (if at all), 4) when the confirmed diagnosis of schwannomatosis was communicated to the patient (if at all) and 5) last follow-up in NF clinic. As diagnostic criteria for schwannomatosis evolved over the study period,^16-18^ a schwannomatosis diagnosis was considered confirmed on the date that patients first fulfilled the diagnostic criteria published at that time. Date of diagnosis communication was taken as the first time a clinician referenced a confirmed schwannomatosis diagnosis in a patient encounter note or other communication with the patient. If precise dates were not available for events prior to patients’ contact with the medical systems, (e.g., ‘Patient first developed pain last March’), we estimated dates using a modified version of the protocol developed for the Cancer Symptom Interval Measure project.^22^ If the medical record contained conflicting information about the timing/nature of medical events and primary source documentation was not available, we privileged data from the medical encounter closest to the original event when choosing which dates to abstract. Full protocols for date estimation and adjudication are available.^23^

**Figure 2.**
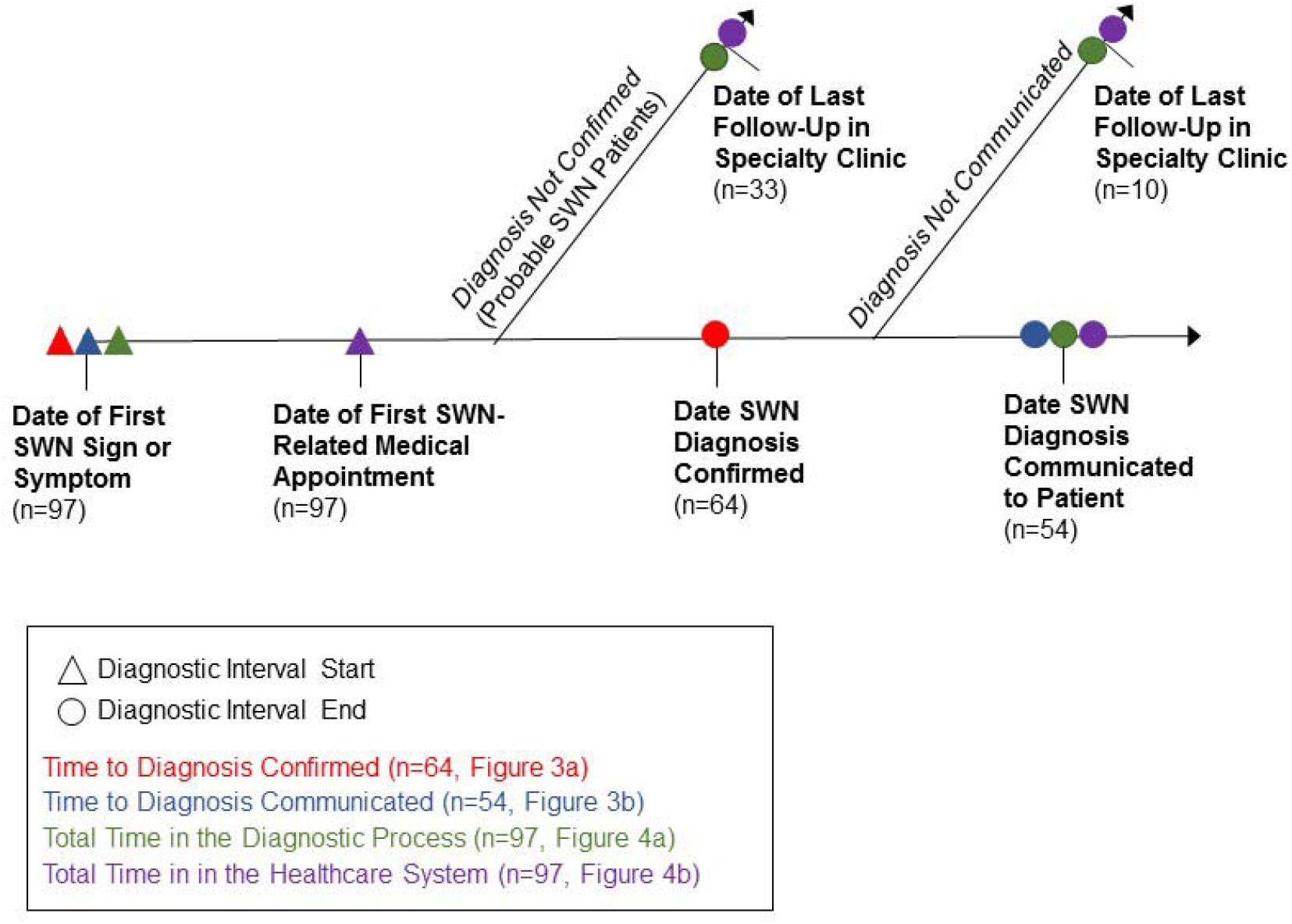
Key Diagnostic Timepoints and Diagnostic Intervals.

#### Diagnostic Errors

We identified cases of misdiagnoses (when another diagnosis was given instead of schwannomatosis) and incomplete diagnoses (when schwannomatosis or another tumor suppressor syndrome should have been considered in the differential diagnosis but was not). Misdiagnoses were defined as “diagnoses that are wrong as judged from the eventual appreciation of more definitive information” (i.e. a clinician attributed a sign or symptom later found to be associated with schwannomatosis to another disease process).^24^ Diagnoses offered as one of multiple options in a differential were not counted as misdiagnoses. All cases of suspected misdiagnosis were reviewed by an expert NF clinician, who was asked to assess whether the preponderance of evidence available in the medical record suggested that the patient was misdiagnosed. This standard is similar to prior studies in which clinical reviewers retrospectively assessed the likelihood of adverse events, and any event with a certainty at or above “more likely than not” or “more than 50/50” was counted as an adverse event.^25,26^ Incomplete diagnoses were defined as situations in which patients received multiple diagnoses of sporadic tumors despite reasonable medical evidence suggesting they may have an underlying tumor suppressor syndrome. This was operationalized as situations in which a patient had two or more nerve sheath tumors (at least one pathologically proven to be a schwannoma), but the patient was not worked up for possible neurofibromatosis 2/schwannomatosis or referred to another provider for such workup.

### Data Analysis

We present descriptive data on patient demographics, clinical characteristics, diagnostic delay, and rates/types of misdiagnosis and incomplete diagnosis. We calculated the time to diagnosis using two endpoints: the date a diagnosis of schwannomatosis was confirmed (i.e. when a patient objectively met diagnostic criteria) and the date a confirmed diagnosis was communicated to the patient (i.e. when the patient was told they had schwannomatosis). Since not all patients had a diagnosis confirmed and communicated to them (either due to lack of communication or because they had probable schwannomatosis that was never confirmed), we also performed a time to event analysis to describe the total time patients spent in the schwannomatosis diagnostic process. We used the date that a diagnosis of schwannomatosis was communicated to the patient as the event, and censored participants without diagnosis confirmation or diagnosis communication on the date of their last NF clinic visit. Finally, to benchmark progress towards the International Rare Diseases Research Consortium’s (IRDIRC) goal of diagnosing all rare disease patients within one year of presentation to medical care,^27^ we also calculated the total time patients spent in the healthcare system (i.e. time from first medical appointment for a sign/symptom of schwannomatosis until communication of confirmed diagnosis or last follow-up in clinic for a probable diagnosis).

We tested the associations between the length of diagnostic delay and clinicodemographic factors (i.e. sex, age at first symptom, inheritance, anatomical extent of disease, initial symptom type, initial symptom frequency, and initial imaging presentation). We grouped patients’ initial signs/symptoms by how specifically they indicated a possible tumor/tumor predisposition syndrome into three categories: asymptomatic (incidental finding or presymptomatic genetic testing); palpable mass with or without other symptoms; and other symptoms only (including pain and neurological symptoms). We used the Mann-Whitney U test (for dichotomous variables) or the Kruskal-Wallis H test with pairwise comparisons via the Dunn-Bonferroni test (for all other categorical variables) to assess for differences in diagnostic delay between patient subgroups. We used Spearman’s rho to assess the strength of associations between continuous variables and diagnostic interval length. All statistical analyses were performed using SPSS Version 24.0 (IBM Corp., Armonk, NY) and all reported p-values are two-tailed.

## Results

### Demographic and Clinical Characteristics

Ninety-seven patients (71 at MGH and 26 at JHH) with schwannomatosis were identified who met inclusion criteria for this study (Table 1). Sixty-four patients had a confirmed diagnosis of schwannomatosis by Plotkin et al. (2013) criteria; 33 patients were classified as having probable schwannomatosis. Overall, 70/97 patients (72.2%) experienced pain, alone or in combination with a palpable mass or neurological symptoms, as their first symptom of schwannomatosis. Ten patients were asymptomatic at the time their health problem was discovered: eight had an incidental imaging finding, one had an incidental finding on physical exam, and one elected to undergo presymptomatic genetic testing after a family member was diagnosed with schwannomatosis.

**Table 1.** Patient Demographic and Clinical Characteristics.

|  | All Schwannomatosis Patients (n=97) | Confirmed Schwannomatosis Patients (n=64) | Probable Schwannomatosis Patients (n=33) |
| --- | --- | --- | --- |
| Female | 49 (50.5%) | 30 (46.9%) | 19 (57.6%) |
| Familial Disease | 15 (15.4%) | 11 (17.2%) | 4 (12%) |
| Anatomically Limited Disease | 20 (20.6%) | 14 (21.9%) | 6 (18%) |
| Age at First Symptom, median (range) | 32.9 years (5.1 – 68.3 years) | 30.8 years (5.1 – 64.1 years) | 39.7 years (12.5 – 68.3 years) |
| First Symptom Type |  |  |  |
| Pain | 51 (52.6%) | 34 (53.1%) | 17 (51.5%) |
| Palpable Mass | 10 (10.3%) | 7 (10.9%) | 3 (9.1%) |
| Neurological Symptoms† | 4 (4.1%) | 0 (0.0%) | 4 (12.1%) |
| Pain and Palpable Mass | 11 (11.3%) | 6 (9.4%) | 5 (15.2%) |
| Pain and Neurological Symptoms | 8 (8.2%) | 7 (10.9%) | 1 (3.0%) |
| Other‡ | 1 (1.0%) | 1 (1.6%) | 0 (0.0%) |
| Unknown | 2 (2.1%) | 2 (3.1%) | 0 (0.0%) |
| N/A: Asymptomatic | 10 (10.3%) | 7 (10.9%) | 3 (9.1%) |
| First Symptom Frequency |  |  |  |
| Constant | 33 (34.0%) | 21 (32.8%) | 12 (36.4%) |
| Intermittent | 29 (29.9%) | 18 (28.1%) | 11 (33.3%) |
| Unknown | 25 (25.8%) | 18 (28.1%) | 7 (21.2%) |
| N/A: Asymptomatic | 10 (10.3%) | 7 (10.9%) | 3 (9.1%) |
| Initial Imaging Workup |  |  |  |
| Single tumor discovered | 54 (55.7%) | 38 (59.4%) | 16 (48.5%) |
| Two or more tumors discovered | 40 (41.2%) | 25 (39.1%) | 15 (45.4%) |
| Unknown | 2 (2.1%) | 1 (1.6%) | 1 (3.0%) |
| N/A – no tumors discovered | 1 (1.0%) | 0 (0%) | 1 (3.0%) |

Footnotes: †Neurological symptoms included numbness, paresthesias, muscle weakness and hypophonia. ‡Other symptom was severe coughing spells due to a schwannoma compressing the esophagus.

Patients consulted a median of five different outpatient healthcare providers (range, 0-20) for diagnosis or treatment of schwannomatosis-related signs and symptoms prior to being diagnosed with confirmed schwannomatosis (or, for people with probable schwannomatosis, their most recent follow-up). There were 554 unique patient/provider pairings documented in the medical record; the specialty of the provider could be determined in 517 (93.3%) cases. Patients most commonly consulted neurosurgeons (n=65, 67%), primary care providers (n=62, 63.9%), orthopedic surgeons (n=42, 43.3%), neurologists (n=40, 41.2%), and general surgeons (n=28, 28.9%).

### Establishing and Communicating a Diagnosis

In the 64 people with confirmed schwannomatosis, median age at diagnosis confirmation was 41.7 years (range, 14.8-81.5 years). Fifty-four of these people (84.4%) had communication of their diagnosis documented in the medical record; median age at diagnosis communication was 41.0 years, range 15.1-81.6 years). Reasons for non-communication in ten patients were as follows. For five patients, the treating physician wanted additional diagnostic evidence beyond what was required by diagnostic criteria to confirm data originally obtained at outside institutions or to obtain additional data to more conclusively rule out possible alternative diagnoses of NF1 and NF2; these data were not obtained before the patients were lost to follow-up or the study period ended. For two patients initially seen prior to publication of the 2013 diagnostic criteria, the treating physician adhered to older diagnostic criteria which the patients did not meet and did not communicate the patient’s updated diagnosis status in subsequent visits after 2013. Finally, no communication was documented in the medical record for three patients who were lost to follow-up.

In the 64 people with confirmed schwannomatosis, the median time from patients’ first sign/symptom of schwannomatosis to the time their diagnosis was confirmed was 6.9 years (range, 0.05 to 47.8 years, 25^th^ – 75^th^ percentile: 3.8 – 16.5 years). At 5, 10, and 20 years from first experience of a health problem related to schwannomatosis, the percentage of people in this group without confirmation of the diagnosis was 67.2%, 37.5%, and 20.3%, respectively (Figure 3a). In the 54 people with confirmed schwannomatosis and documented communication of this diagnosis to the patient, median time to diagnosis communicated was 7.3 years (range, 0.2 to 48.0 years; 25^th^ – 75^th^ percentile: 4.5 – 15.8 years). At 5, 10, and 20 years from first experience of a health problem related to schwannomatosis, the percentage of people in this group without documented communication of the diagnosis was 68.5%, 35.2%, and 20.4%, respectively (Figure 3b).

**Figure 3.**
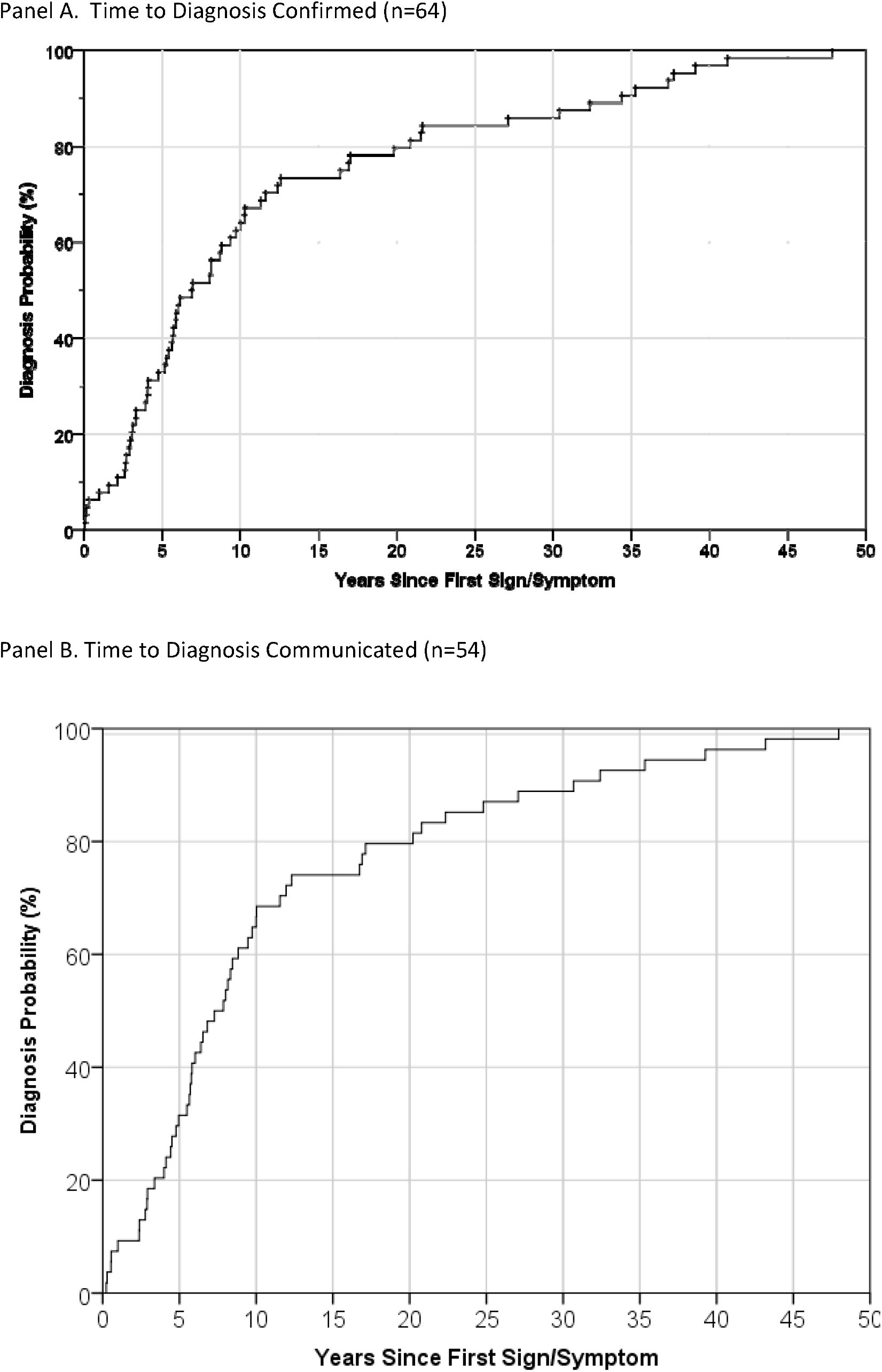
Time to Diagnosis in People with Confirmed Schwannomatosis.

Since not all patients had a confirmed and communicated diagnosis of schwannomatosis, we also performed a time-to-event analysis of total time to diagnosis in all 97 patients, censoring people without a confirmed diagnosis or without documented communication of a confirmed diagnosis at the time of their last follow-up in NF clinic (Figure 4a). Using this method, which accounts for the time spent in the diagnostic process by patients who have not yet had a diagnosis confirmed and/or communicated, the median total time spent in the diagnostic process was 16.7 years (95% CI, 7.5 – 26.0 years). At 5, 10, and 20 years from first symptom, the percentage of people without documented confirmation and communication of the diagnosis was 81.5%, 58.9%, and 45.4%, respectively. To isolate the proportion of this time that was spent in the healthcare system, we looked at the same hybrid endpoint (date of confirmed diagnosis communication or date of last follow-up in NF clinic), but beginning at the date of patients’ first medical appointment for a sign/symptom of schwannomatosis (Figure 4b). The median time spent in the healthcare system seeking diagnosis for all 97 patients was 9.8 years (95% CI, 3.5 – 16.2 years). Only 7.3% of patients had documented diagnosis communication with one year of presenting to care.

**Figure 4.**
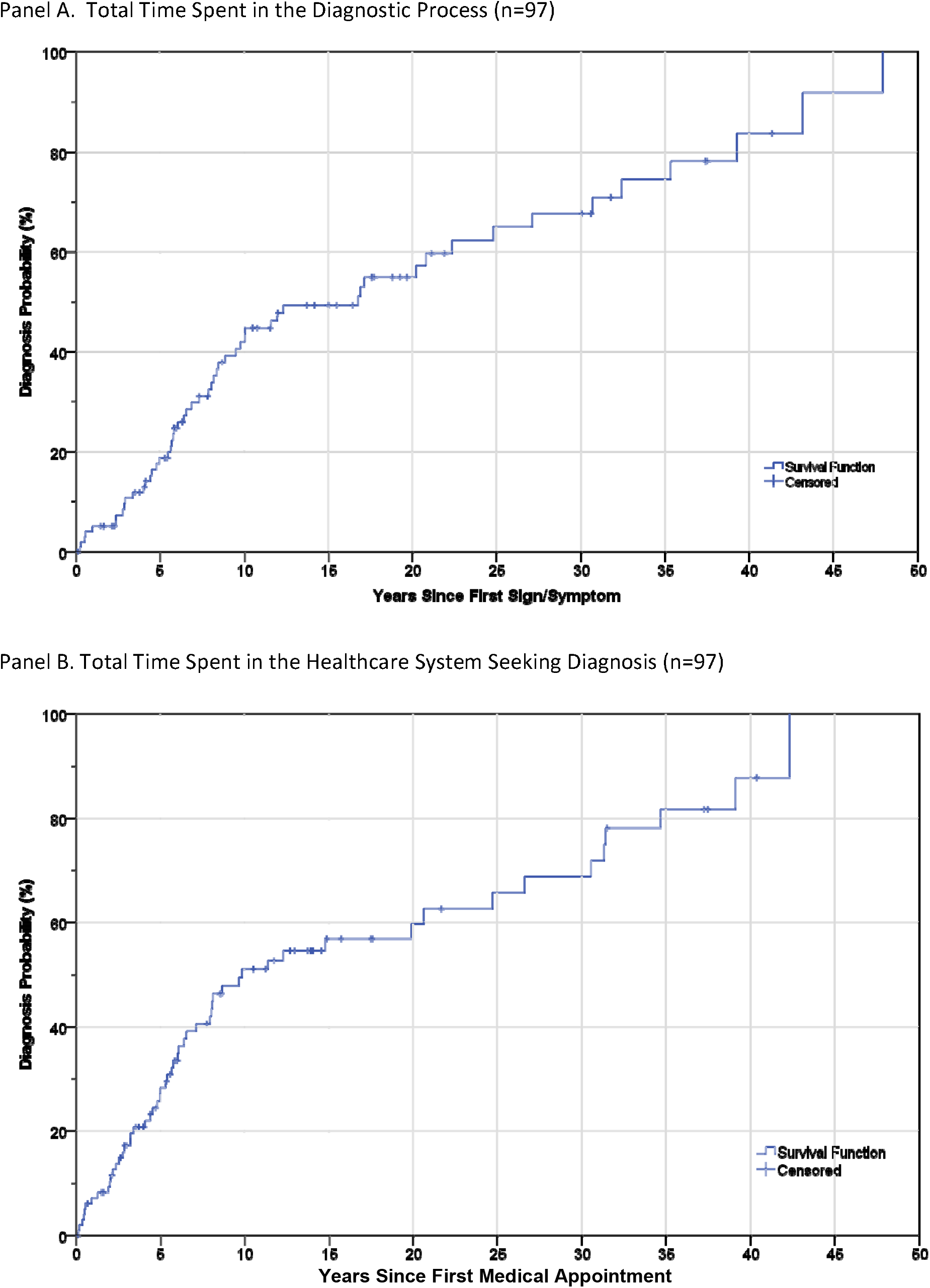
Kaplan Meier Curve of Time Spent in the Diagnostic Process for Entire Cohort.

### Predictors of Diagnostic Delay

We examined the influence of various factors on the total time to diagnosis communicated (n=54) and total time spent in the diagnostic process (n=97), (Table 2). There was no statistically significant effect of patient sex, inheritance pattern, or anatomical extent of disease on either diagnostic time interval, although women trended towards shorter diagnostic intervals. People who experienced their first symptom at younger ages had significantly longer diagnostic intervals (Rho = -0.31, p=0.02 for time to diagnosis communicated; Rho = -0.39 p<0.001 for total time in the diagnostic process.)

**Table 2.** Association between Clinical and Demographic Characteristics and Time to Schwannomatosis Diagnosis.

|  | Time to Diagnosis Communicated (n=54) | Total Time in the Diagnostic Process† (n=97) |
| --- | --- | --- |
| *Sex*<br>Female vs. Male | 5.77 vs. 9.87<br>U=257, p=0.06 | 6.29 vs 10.01<br>U=934.5, p=0.08 |
| *Inheritance Pattern*<br>Sporadic vs. Familial | 8.00 vs. 5.87<br>U=151, p=0.44 | 9.05 vs. 5.86<br>U=484.5, p=0.193 |
| *Extent of Disease*<br>Full vs. Anatomically Limited | 6.83 vs. 8.45<br>U=198, p=0.41 | 7.86 vs 9.23<br>U=704, p=0.56 |
| Age at First Symptom | Rho = -0.31<br>**p=0.02** | Rho = -0.39<br>**p<0.001** |
| *Initial Symptom Type‡*<br>Asymptomatic vs. Palpable Mass vs. Other Symptoms | 4.77 vs. 3.46 vs. 8.32<br>H(2)=8.44, **p=0.02** | 4.64 vs. 5.86 vs. 9.87<br>H(2)=10.79,<br>**p=0.005** |
| *Initial Symptom Frequency*<br>Asymptomatic vs. Constant vs. Intermittent vs. Unknown | 4.77 vs. 4.00 vs. 8.58 vs. 10.98<br>H(3)=14.69, **p=0.002** | 4.64 vs. 5.84 vs. 9.74 vs. 11.95<br>H(3)=16.79,<br>**p=0.001** |
| *Initial Imaging Presentation§*<br>Single Tumor vs. Multiple Tumors | 8.32 vs. 5.63<br>U=810.5, **p=0.04** | 9.19 vs. 6.61<br>U=229, **p=0.04** |

**Table Legend:** For categorical variables, median diagnostic interval length is reported in years. Test statistics and p-values are reported using Mann Whitney U (for dichotomous variables), Kruskal-Wallis H test (for other categorical variables), and Spearman's Rho (for continuous variables).
†Time spent in the diagnostic process extends to the time a diagnosis was communicated, or if that did not occur, the time of the person's last follow-up in NF clinic.
‡ "Palpable mass" includes mass with or without additional symptoms; "other symptoms" includes pain, neurological symptoms or severe coughing spells only; and 2 people with unknown symptoms were excluded from analysis.
§ Three people with either zero or an unknown number of tumors were excluded from analysis.

There were significant differences in diagnostic interval length based on initial symptom type (Table 2, p=0.02 for diagnosis communicated and p=0.005 for total time in diagnostic process). Post-hoc pairwise Dunn-Bonferroni comparisons showed that patients with pain/neurological symptoms had significantly longer time to diagnosis communication than those with a palpable mass (p=0.03) and significantly longer time spent in the diagnostic process than asymptomatic patients (p=0.01). Initial symptom frequency was also significantly associated with the length of both diagnostic intervals (Table 2, p=0.002 for diagnosis communicated and p=0.001 for total time in diagnostic process). Post-hoc pairwise Dunn-Bonferroni tests indicated that patients with intermittent symptoms had significantly longer time to diagnosis communicated than patients with constant symptoms (p=0.02) and more time spent in the diagnostic process than asymptomatic patients (p=0.02). Finally, patients in whom initial imaging workup revealed multiple suspected nerve sheath tumors had significantly shorter diagnostic intervals than patients who had a single nerve sheath tumor (Table 2, p=0.04 for both time to diagnosis communicated and total time spend in the diagnostic process).

### Diagnostic Errors

#### Misdiagnosis

Thirty-five patients (36.1%, 23 confirmed and 12 probable diagnoses) had one or more schwannomatosis-related signs/symptoms that were likely misdiagnosed based on subsequent expert clinical review. Misdiagnoses fell into three, non-mutually exclusive categories: patients’ underlying genetic disorder being misdiagnosed as another genetic condition; tumors being mis-classified based on physical exam, imaging, or pathological characteristics; and pain etiology being misattributed to a more common disease process.

Eighteen patients (18.6%) had their overall genetic condition misdiagnosed: 13 with NF1, four with NF2, and one with both NF1 and NF2 (by different providers). Eleven patients (11.3%) had a nerve sheath tumor misdiagnosed. Six patients had resected masses which were originally classified as neurofibromas by pathologists at outside hospitals, but were re-classified as schwannoma (five cases) or hybrid nerve sheath tumor (one case) upon review by an expert NF pathologist. Two of these patients also had additional, later schwannomas misdiagnosed as NF1-related lesions (malignant peripheral nerve sheath tumor in one case and gastrointestinal stromal tumor in one case). Five patients had masses that were diagnosed as enlarged lymph nodes, cysts, or vascular lesions based on the presence of radiological and/or physical exam characteristics alone, but were later resected with pathology of schwannoma.

Sixteen patients (16.5%) had the etiology of their pain likely misdiagnosed. Pain was most commonly misattributed to neuromuscular or orthopedic issues such as degenerative disc disease, carpal tunnel syndrome, sciatica, bursitis, plantar fasciitis, tendonitis, and arthritis. Less commonly, pain was attributed to an injury (such as abdominal strain, after-effects of childbirth, and a rotator cuff injury) or to psychological issues (such as psychosomatic pain). For most patients, later identification and removal of a schwannoma led to pain reduction, lending credence to the idea that alternate diagnoses, even if they existed as comorbidities, were not the primary source of patients’ pain.

#### Incomplete diagnosis

We also observed 19 patients (19.6%, 13 confirmed and six probable) where an underlying genetic syndrome should have been investigated earlier as a potential diagnosis. In 13/19 cases, there was sufficient detail to suggest physician error (i.e. documentation showed that an individual physician had knowledge of the patient’s multiple schwannomas and neither began workup for potential NF2/schwannomatosis nor referred the patient to a specialist for further workup). In the remaining 6/19 cases, it was unknown whether a single physician had knowledge of the patient’s multiple schwannomas, and as such the error could have been due to system issues (for example, a failure to transmit documentation of patient’s prior schwannoma between hospitals, such that a physician was unaware the patient had multiple tumors and met criteria for further NF2/schwannomatosis workup).

## Discussion

In this study, we reviewed the medical records of 97 people with confirmed or probable schwannomatosis to assess diagnostic accuracy and timeliness in patients eventually seen in specialized, tertiary care clinics. Our research shows that prolonged times to schwannomatosis diagnosis are common, with a median time to diagnosis communication of 7.3 years in confirmed patients, 20% of whom were not diagnosed until ≥20 years after their first symptom. These estimates are even greater when considering the large number of patients who have not yet received a confirmed diagnosis of schwannomatosis. In a time-to-event analysis accounting for the time patients with probable schwannomatosis have already spent undergoing diagnostic workup, we found that the median time spent in the diagnostic process for all patients was 16.7 years. While there is no clearly accepted dividing line between what constitutes a timely or delayed diagnosis of schwannomatosis, it is clear that a significant number of patients face extended diagnostic intervals and substantial room for improvement exists. Only 7.3% of patients met the IRDIRC goal of diagnosing all rare disease patients within one year of presentation to medical care.^27^

We identified several disease- and patient-related factors associated with time to diagnosis, which at least in part were related to patients’ initial symptom appraisal and help-seeking. Asymptomatic patients with incidentally found tumors or who electively underwent presymptomatic screening for schwannomatosis due to an established family history of the disorder were diagnosed more quickly. Conceptually, these patients were able to skip the typical delay associated with recognizing one has a health problem, deciding to seek care, and obtaining an appointment (Figure 1). Conversely, patients with intermittent or less specific tumor symptoms (i.e. pain and/or neurological symptoms only) had prolonged times to diagnosis. Possibly patients did not perceive these symptoms as sufficiently abnormal or as bothersome enough to warrant medical attention, leading to what Andersen, Cacioppo and Roberts (1995) called appraisal and illness delays in receiving care.^28^ Finally, younger patients also had delayed diagnoses. Previous research has shown that younger adults are more likely to avoid seeing a doctor even when they suspect medical care is necessary, perhaps due to increased belief that symptoms will improve on their own and/or lower likelihood of having an existing usual source of care.^29,30^ Given that adolescents and young adults with other tumor types have also been shown to experience longer diagnostic intervals than either older adults or young children,^31,32^ further efforts are needed to effectively diagnose schwannomatosis in young adults.

Misdiagnosis of schwannomatosis-related signs and symptoms were also common, with more than one-third of patients likely experiencing a misdiagnosis. Surprisingly, misdiagnoses of NF1 (in which patients are prone to multiple neurofibromas) were more common than misdiagnoses of NF2 (which like schwannomatosis, predisposes patients to develop multiple schwannomas). NF1 is much more prevalent than NF2 and schwannomatosis, so physicians may be more familiar with this disorder.^8,33^ NF1 misdiagnoses were also often related to prior pathological misdiagnoses of schwannomas or hybrid nerve sheath tumors as ‘neurofibromas’. Hybrid nerve sheath tumors, which display features of both neurofibroma and schwannoma,^34,35^ are common in schwannomatosis.^36^ Careful pathological review of these tumors, as well as the subtypes of schwannomas most likely to be misdiagnosed (e.g. cellular schwannomas and those with myxoid change), may help improve diagnostic accuracy.^37^ Clinically, expert pathologic re-review with newer molecular tests^38^ is likely indicated in people who have prior pathology findings of both neurofibromas and schwannoma (to appropriately classify them as either NF1 spectrum or NF2/Schwannomatosis spectrum), and in people with multiple ‘neurofibromas’ who do not display other clinical features of NF1 (as they may be misdiagnosed schwannomatosis patients).

Other NF1 and NF2 misdiagnoses point to the difficulty of diagnosing genetic conditions, which may present in a segmental pattern – that is, affecting only a portion of the body – or with different symptoms/signs based on one’s unique genetic mutation. Even when treating physicians noted that patients lacked the characteristic skin findings of NF1 (café au lait spots, skin fold freckling, and cutaneous neurofibromas), patients could still be misdiagnosed with mosaic NF1 or a variant of NF1 that primarily causes spinal tumors under the assumption that genetic features explained patients’ lack of skin findings. Similarly, even in patients without vestibular schwannoma (a defining characteristic of NF2), patients could be assumed to have mosaic NF2, which indeed displays extensive phenotypic overlap with schwannomatosis.^8,19^ More widespread use of genetic testing may be the only reliable way to distinguish these possibilities, supporting recent recommendations by an international consensus panel for routine use of diagnostic genetic testing for all cases of NF2 and schwannomatosis.^39^

Finally, clinicians’ lack of familiarity with NF1, NF2 and schwannomatosis likely contributed to some incomplete diagnoses. One-fifth of people with multiple suspected tumors radiologically - at least one of which had been pathologically confirmed as a schwannoma - were not evaluated for possible genetic predispositions or referred to another provider for such workup. Given how rarely multiple nerve sheath tumors occur in the absence of a tumor predisposition syndrome, appearance of a second potential schwannoma should serve a trigger symptom to initiate neurogenetic consultation. To avoid solely relying on clinicians to remember and act on this trigger symptom, thoughtful use of electronic medical record alerts may be helpful. For example, patients with pathology records showing more than one nerve sheath tumor and/or patients with radiology reports mentioning multiple potential nerve sheath tumors could be flagged for neurogenetic workup. Similar electronic trigger algorithms have already been developed to improve delays in follow-up after abnormal lung CT and mammography findings^40,41^ and have been shown in randomized controlled trials to reduce diagnostic delay in colorectal and prostate cancer.^42^

### Study Limitations

Completeness of medical documentation varied from patient to patient, particularly regarding documentation from providers outside the MGH and JHH hospital networks. This limited our ascertainment of precise dates for the first medical appointment for some patients, as well our ability to catch misdiagnoses or incomplete diagnoses that happened early in the diagnostic process. Retrospective ascertainment of misdiagnosis is inherently difficult; to make our assessment as robust as possible, we based our evidentiary standard on prior medical studies^25,26^ and explicitly excluded suspected misdiagnoses that could not be corroborated based on a “preponderance of evidence”. While prioritizing diagnostic communication as our primary endpoint aligned our analysis with the NAM model of diagnosis, there are also difficulties in assessing communication using medical records. Phone calls and electronic communications were less well documented than in-person visits, and questions remain regarding the degree to which clinicians’ verbal communication and patients’ understanding of their diagnosis match what is documented in medical records. Thus, our analysis may best be considered a proxy measure of true communication and understanding. Finally, our results may not be generalizable to all schwannomatosis patients, since our sample includes only those people who eventually attended dedicated NF clinics at large academic medical centers. While the delays and errors that patients at tertiary care clinics have experienced may not be identical to those of people who are still undiagnosed or managed outside specialty center, the common problems identified in the present study nonetheless represent important targets for process improvement.

## Conclusions

Our study is the first comprehensive assessment of diagnostic performance in people with schwannomatosis seen at specialized, U.S.-based clinics. By assessing key dimensions of diagnostic error identified by the National Academy of Medicine — timeliness and accuracy in establishing a diagnosis and communicating it to the patient^6^ — we have established benchmarks for future quality improvement efforts in the areas most impactful to patient experience. Our work highlights several target areas for improving diagnosis of schwannomatosis, including increasing clinician awareness of schwannomatosis and how it differs from other types of NF; instituting more widespread genetic testing for potential schwannomatosis cases and expert review of selected pathology findings; and testing new methods to automatically flag patients for genetic evaluation based on imaging and/or pathology findings. Future research in schwannomatosis should prioritize piloting interventions with broad potential to improve diagnosis across rare disorders, including health IT interventions like electronic trigger algorithms, real-time diagnostic decision support, and health information exchanges.^43-46^ These ambitious, systemwide efforts may be best poised to improve the clinical management of people with rare conditions and ensure that all patients and their family members have access to appropriate genetic and reproductive counseling.

## Data Availability

All data produced in the present study are available upon reasonable request to the authors.

## Acknowledgments

Funding for this research was provided by a Children’s Tumor Foundation Young Investigator Award to VM and the Program in Cancer Outcomes Research Training (National Cancer Institute R25CA92203).

## Notes

Conflicts of Interest: Vanessa Merker reports consulting income from the Neurofibromatosis Network. Justin Jordan reports consulting income from Navio Theragnostics, CereXis, Recursion, Health2047; <1% stock ownership in Navio Theragnostics; Royalties from Elsevier. Jaishri Blakeley has served as a paid consultant for Astella Pharma, Abbvie, TriAct/BiPar Sciences and has had research support from GlaxoSmithKlein, Bristol Meyer Squibb, and AstraZeneca. Scott Plotkin is co-founder of NFlection Therapeutics and NF2 Therapeutics; serves on the Scientific Advisory Board and has stock in SonALAsense; and consults for Akouos.

### Competing Interest Statement

Vanessa Merker reports consulting income from the Neurofibromatosis Network. Justin Jordan reports consulting income from Navio Theragnostics, CereXis, Recursion, Health2047; <1% stock ownership in Navio Theragnostics; Royalties from Elsevier. Jaishri Blakeley has served as a paid consultant for Astella Pharma, Abbvie, TriAct/BiPar Sciences and has had research support from GlaxoSmithKlein, Bristol Meyer Squibb, and AstraZeneca. Scott Plotkin is co-founder of NFlection Therapeutics and NF2 Therapeutics; serves on the Scientific Advisory Board and has stock in SonALAsense; and consults for Akouos.

### Funding Statement

This study was funded by Childrens Tumor Foundation Young Investigator Award 2015-01-005 and the Program in Cancer Outcomes Research Training (National Cancer Institute R25CA92203).

### Author Declarations

The Institutional Review Boards at Mass General Brigham, Johns Hopkins Medicine, and Boston University Medical Center gave ethical approval for this work. A waiver of informed consent was granted for this retrospective medical record review.

### Summary of Updates

Minor wording changes throughout manuscript text and inclusion of a revised Figure 2.

